# Speech markers of depression: cross-sectional discrimination, longitudinal association, and treatment response prediction

**DOI:** 10.64898/2026.10.05.26364692

**Authors:** Khatiya C Moon, Leily M Behbehani, Anil K Malhotra, Michael J Spilka, Sunny X Tang

## Abstract

**Objective:** To identify speech features distinguishing adults with depression from healthy controls, evaluate whether these features track with depression severity during naturalistic treatment, and assess whether baseline speech features predict treatment response.

**Design:** Cross-sectional group comparison (Objective A) followed by longitudinal evaluation in an independent sample (Objectives B and C). Candidate features surviving false discovery rate (FDR) correction in the cross-sectional analysis were carried forward to longitudinal and predictive analyses.

**Setting:** A community mental health center in New York.

**Participants:** Dataset I (cross-sectional): 82 adults with chart-diagnosed depressive disorders and 80 healthy controls (N=162), recruited via convenience sampling. Dataset II (longitudinal): 85 adults with depressive disorders followed for up to four timepoints over a mean of 10±3.5 weeks weeks during naturalistic treatment; 63 with baseline PHQ-9 >=10 and at least one follow-up were included in prediction analyses. Recruitment occurred under protocols approved between 2020 and 2021.

**Interventions:** None; participants received naturalistic clinical care.

**Outcome Measures:** Cross-sectional: diagnostic group membership (depressive disorder vs. healthy control). Longitudinal: Patient Health Questionnaire-9 (PHQ-9) total score. Predictive: treatment response, defined as >=50% PHQ-9 reduction from baseline to last follow-up.

**Results:** Across three speech tasks (journaling, picture description, phonemic fluency), 11 voice quality features, six shimmer variants and five harmonics-to-noise ratio (HNR) variants, survived FDR correction (all q<.05) in all tasks, with higher shimmer and lower HNR in the depressed group (largest adjusted effect beta=1.89, q<.001). In longitudinal analysis, one phonemic fluency voice quality feature was associated with PHQ-9 at FDR<.05 (maximum HNR beta=-1.43, SE=0.41, FDR=.006), with the remaining ten consistent in direction at FDR<.10; no speech x time interactions were significant. Sex moderated voice quality-depression associations in picture description and phonemic fluency. For treatment response prediction, phonemic fluency voice quality features achieved cross-validated AUC=0.727 [95% CI: 0.662, 0.792], comparable to clinical features alone (AUC=0.700-0.715); the combined model across all tasks and clinical features achieved AUC=0.730 [0.639, 0.820]. No pairwise AUC comparisons between predictor configurations reached significance after correction (all corrected p>.54).

**Conclusions:** Voice quality features, particularly shimmer and HNR, were the most consistent speech correlates of depression across cross-sectional, longitudinal, and predictive analyses and across multiple speech tasks. These features are computationally simple to extract from brief recordings and showed preliminary predictive utility comparable to standard clinical variables. However, the modest sample sizes, chart-based diagnoses and heterogeneous treatment exposures preclude clinical application at this stage. Larger, prospectively designed studies with standardized recording protocols, structured diagnostic assessment, and recruitment at treatment initiation are needed before voice quality features can be recommended as clinical biomarkers.

## Introduction

Depressive disorders are among the largest contributors to global disability and mortality (Zhang 2025, Zhou 2025, Dong 2019, Cai 2021, Cai 2021). Both diagnosis and treatment planning of depression rely primarily on clinician interview and subjective patient report, despite that clinician skill and approach may vary widely, and patient report may be compromised by illness-related effects on insight and social engagement. Half of patients with depression that present to care are not diagnosed at their initial encounter (Garcia 2022). Even when treated, no objective biomarkers exist to guide treatment recommendations and most patients do not receive treatment that conforms to best practice standards (Simon 2024). Even with the best treatment, only 36% of patients achieve remission after an initial antidepressant trial (Rush 2006). Simultaneously, most first-line antidepressants (e.g. selective serotonin reuptake inhibitors) take multiple weeks or months to take full effect. Thus, patients and clinicians undergo a months-long process of trial-and-error based treatment, resulting in delayed recovery and ongoing disability. Objective, scalable biomarkers are needed to both help diagnose depression as well as predict treatment response and treatment trajectory.

Decades of efforts to identify objective biomarkers of depression and its treatment response have spanned inflammatory, neurotrophic, neuroendocrine, genomic, and neuroimaging domains, yet none has entered routine clinical use (Zallar 2026). The earliest modern conceptualizations of depression described it as a disorder producing slowed, sparse, and monotonous speech, setting in motion a line of research investigating speech as a candidate biomarker (Kendler 2017). Speech is a complex output combining vocal acoustic and linguistic features that reflect cognitive, physiological, and emotional states. Compared to other candidate biomarkers that require magnetic resonance imaging, phlebotomy, or electroencephalography (EEG), speech requires no specialized space, equipment, or personnel to collect. Speech samples can be collected automatically and remotely using smartphones or web apps. Analysis of speech can be conducted through automated pipelines. In this way, speech is highly scalable candidate biomarker and feasible for repeated routine clinical use as well as in low-resource and/or remote settings.

Since the 1970s a body of literature has emerged identifying speech timing features (increased pause duration, length of speech, response time) from recorded samples as features that distinguish depressed individuals from healthy controls (Yamamoto 2020), largely validating the initial observations of depressed patients made by Kraeplin and other early theorists. However, computational advances have enabled extraction of a far broader range of speech features from recorded samples. Contemporary automated pipelines can quantify not only temporal characteristics (e.g., speech rate, pause duration) but also acoustic and voice quality properties (e.g., fundamental frequency, shimmer, harmonics-to-noise ratio), as well as linguistic features: morphosyntactic complexity, semantic coherence, and lexical diversity. Together, high-dimensional feature sets capture aspects of speech production that span motor, cognitive, and affective processes (Yawatkar, 2025).

Cross-sectional studies comparing depressed individuals to healthy controls have identified differences across many of these domains (Maran 2025, Mao 2023, Briganti 2025). Systematic reviews and meta-analyses report reduced speech rate, increased pause duration, lower pitch variability, and altered voice quality in depression, though effect sizes vary across studies and feature definitions are not always consistent (Cantor-Cutiva 2025, Mao 2023). Far fewer studies have examined speech features longitudinally either as markers that track with symptom change during treatment or as baseline predictors of treatment response (Mundt 2012; Mundt 2007; Yamamoto 2020; Briganti 2025). Mundt et al. (2007) first demonstrated in an observational study of voice acoustic features extracted through telephone interactive voice recordings that greater pitch variability, reduced pause duration, and faster speech rate differentiated responders and non-responders, and Mundt et al. (2012) subsequently replicated these findings in a larger randomized, placebo-controlled sample. These studies examined a small set of timing and prosodic features, leaving open whether interrogation of a broader acoustic and linguistic feature space could identify additional or more robust markers. Moreover, Mundt’s models were based on speech change during treatment rather than baseline speech, leaving unanswered whether pre-treatment speech features can predict who will respond before treatment begins. Finally, aside from the Mundt papers, few studies have tested whether features identified as discriminative in one sample replicate in independent cohorts, leaving open whether cross-sectional group differences reflect reproducible, clinically relevant signals or sample-specific findings.

### The Present Study

The present study addressed these gaps through three objectives applied across two independent samples. In a cross-sectional sample we identified speech features that distinguished adults with depressive disorders from healthy controls (Objective A). Features surviving correction for multiple comparisons were then carried forward to an independent longitudinal sample of adults with depression undergoing naturalistic treatment where we evaluated whether these features tracked with depression severity over time (Objective B) and whether baseline features predicted treatment response (Objective C).

## Methods

### Recruitment, data acquisition, extraction, and processing

#### Ethical considerations

All participants (Table 1) provided written informed consent, and all study protocols were approved by the institutional review board at the Feinstein Institutes for Medical Research (#20-0468, # 20-0360, and # 21-0259).

**Table 1.**
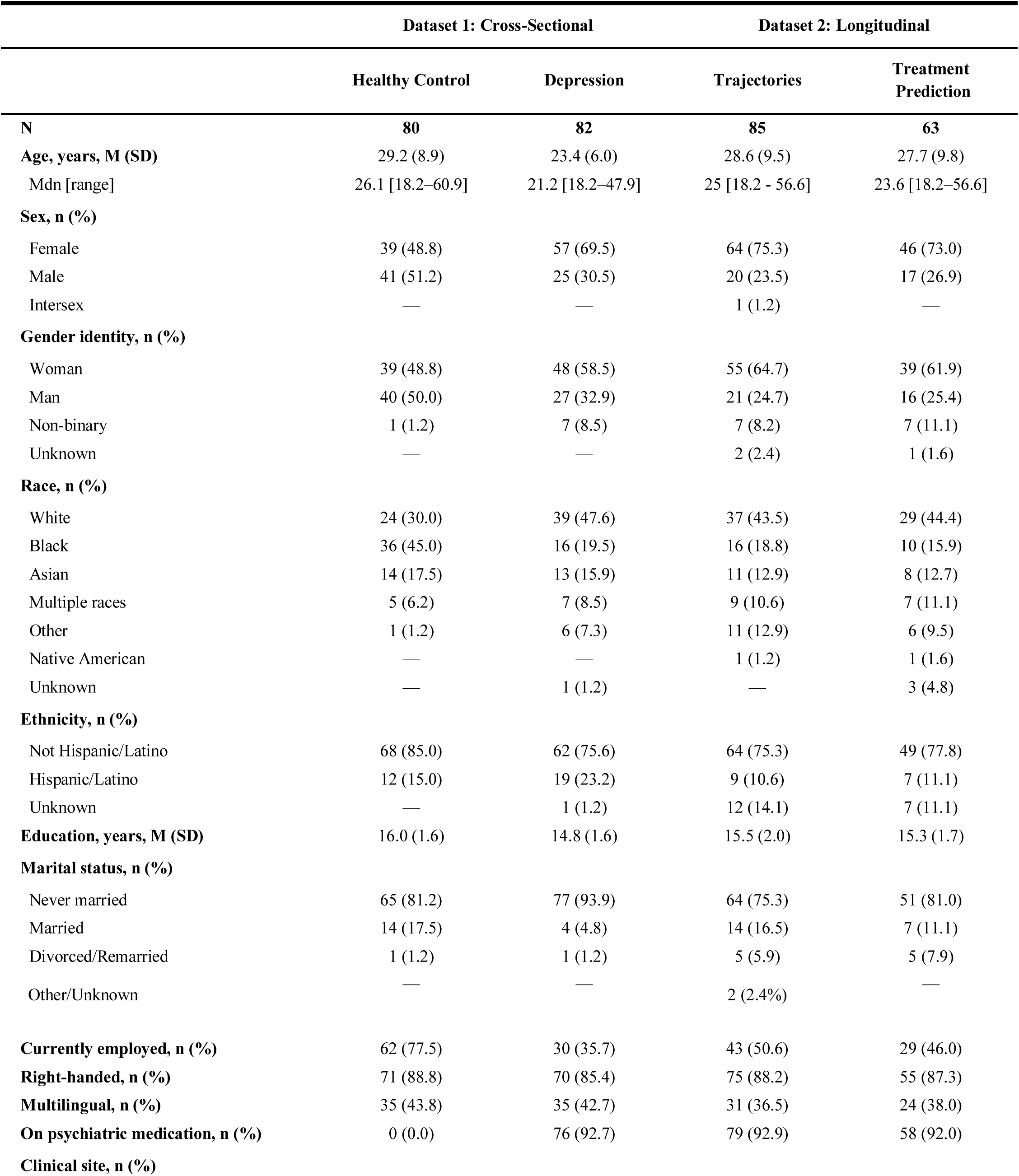
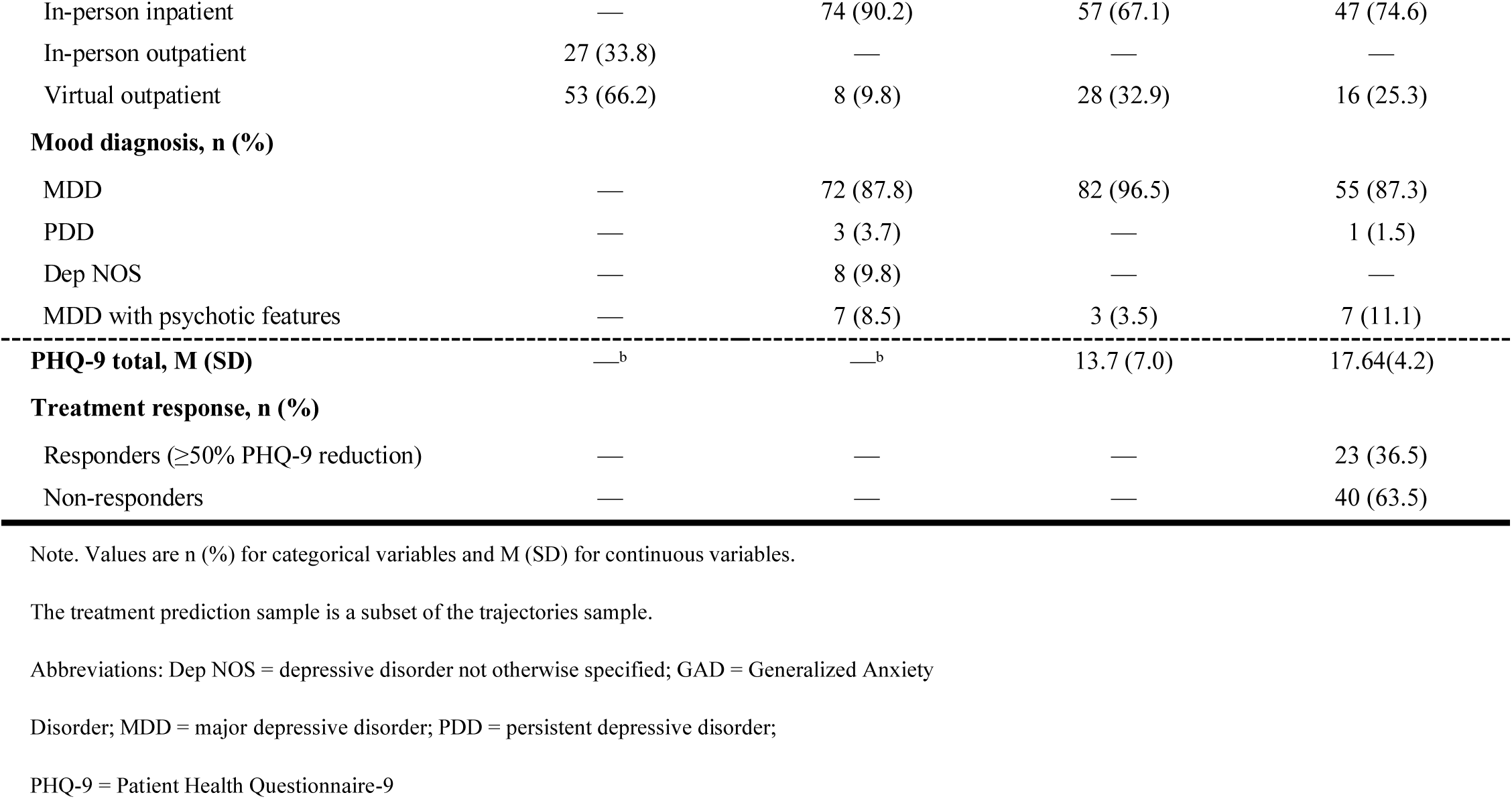
Participant Characteristics.

|  | Dataset 1: Cross-Sectional |  | Dataset 2: Longitudinal |  |
| --- | --- | --- | --- | --- |
|  | Healthy Control | Depression | Trajectories | Treatment Prediction |
| N | 80 | 82 | 85 | 63 |
| Age, years, M (SD) | 29.2 (8.9) | 23.4 (6.0) | 28.6 (9.5) | 27.7 (9.8) |
| Mdn [range] | 26.1 [18.2–60.9] | 21.2 [18.2–47.9] | 25 [18.2 - 56.6] | 23.6 [18.2–56.6] |
| Sex, n (%) |  |  |  |  |
| Female | 39 (48.8) | 57 (69.5) | 64 (75.3) | 46 (73.0) |
| Male | 41 (51.2) | 25 (30.5) | 20 (23.5) | 17 (26.9) |
| Intersex | — | — | 1 (1.2) | — |
| Gender identity, n (%) |  |  |  |  |
| Woman | 39 (48.8) | 48 (58.5) | 55 (64.7) | 39 (61.9) |
| Man | 40 (50.0) | 27 (32.9) | 21 (24.7) | 16 (25.4) |
| Non-binary | 1 (1.2) | 7 (8.5) | 7 (8.2) | 7 (11.1) |
| Unknown | — | — | 2 (2.4) | 1 (1.6) |
| Race, n (%) |  |  |  |  |
| White | 24 (30.0) | 39 (47.6) | 37 (43.5) | 29 (44.4) |
| Black | 36 (45.0) | 16 (19.5) | 16 (18.8) | 10 (15.9) |
| Asian | 14 (17.5) | 13 (15.9) | 11 (12.9) | 8 (12.7) |
| Multiple races | 5 (6.2) | 7 (8.5) | 9 (10.6) | 7 (11.1) |
| Other | 1 (1.2) | 6 (7.3) | 11 (12.9) | 6 (9.5) |
| Native American | — | — | 1 (1.2) | 1 (1.6) |
| Unknown | — | 1 (1.2) | — | 3 (4.8) |
| Ethnicity, n (%) |  |  |  |  |
| Not Hispanic/Latino | 68 (85.0) | 62 (75.6) | 64 (75.3) | 49 (77.8) |
| Hispanic/Latino | 12 (15.0) | 19 (23.2) | 9 (10.6) | 7 (11.1) |
| Unknown | — | 1 (1.2) | 12 (14.1) | 7 (11.1) |
| Education, years, M (SD) | 16.0 (1.6) | 14.8 (1.6) | 15.5 (2.0) | 15.3 (1.7) |
| Marital status, n (%) |  |  |  |  |
| Never married | 65 (81.2) | 77 (93.9) | 64 (75.3) | 51 (81.0) |
| Married | 14 (17.5) | 4 (4.8) | 14 (16.5) | 7 (11.1) |
| Divorced/Remarried | 1 (1.2) | 1 (1.2) | 5 (5.9) | 5 (7.9) |
| Other/Unknown | — | — | 2 (2.4%) | — |
| Currently employed, n (%) | 62 (77.5) | 30 (35.7) | 43 (50.6) | 29 (46.0) |
| Right-handed, n (%) | 71 (88.8) | 70 (85.4) | 75 (88.2) | 55 (87.3) |
| Multilingual, n (%) | 35 (43.8) | 35 (42.7) | 31 (36.5) | 24 (38.0) |
| On psychiatric medication, n (%) | 0 (0.0) | 76 (92.7) | 79 (92.9) | 58 (92.0) |
| Clinical site, n (%) |  |  |  |  |
| In-person inpatient | — | 74 (90.2) | 57 (67.1) | 47 (74.6) |
| In-person outpatient | 27 (33.8) | — | — | — |
| Virtual outpatient | 53 (66.2) | 8 (9.8) | 28 (32.9) | 16 (25.3) |
| **Mood diagnosis, n (%)** |  |  |  |  |
| MDD | — | 72 (87.8) | 82 (96.5) | 55 (87.3) |
| PDD | — | 3 (3.7) | — | 1 (1.5) |
| Dep NOS | — | 8 (9.8) | — | — |
| MDD with psychotic features | — | 7 (8.5) | 3 (3.5) | 7 (11.1) |
| **PHQ-9 total, M (SD)** | —<sup>b</sup> | —<sup>b</sup> | 13.7 (7.0) | 17.64(4.2) |
| **Treatment response, n (%)** |  |  |  |  |
| Responders (≥50% PHQ-9 reduction) | — | — | — | 23 (36.5) |
| Non-responders | — | — | — | 40 (63.5) |

Note. Values are n (%) for categorical variables and M (SD) for continuous variables.
The treatment prediction sample is a subset of the trajectories sample.
Abbreviations: Dep NOS = depressive disorder not otherwise specified; GAD = Generalized Anxiety
Disorder; MDD = major depressive disorder; PDD = persistent depressive disorder;
PHQ-9 = Patient Health Questionnaire-9

#### Recruitment

The nonoverlapping datasets described below were recruited from Zucker Hillside Hospital (Glen Oaks, NY), a community mental health center with inpatient and outpatient facilities, using convenience sampling of patients cleared by their treatment teams for participation. Diagnosis was determined from the clinical chart in both datasets.

#### Speech Tasks

Both datasets elicited three speech tasks: journaling (open-ended narrative responses), picture description, and phonemic fluency (F-letter word generation for one minute) (Ferreria, 2024; Patterson 2011). Additional tasks which were not reproduced across datasets were not included for analysis (i.e., paragraph reading and semantic fluency in Dataset I, sustained phonation in Dataset II). Task stimuli differed in some respects between datasets. For journaling, Dataset I used the prompts “Tell me about yourself” and “How have you been spending your time recently?”, while Dataset II used “How have you been spending your time recently?” and “Briefly describe all the positive things you expect to happen this week”. For picture description, both datasets used three visual stimuli per session; Dataset I stimuli included images derived from the Thematic Apperception Test (Morgan & Murray, 1935) and the Rorschach Test (Rorschach, 1942), and Cookie Theft (Roth, 2011).

#### Acquisition Modality

Speech assessments took approximately 10–15 minutes to complete. For Dataset I, participants completed tasks on iPads via the Winterlight iOS application during sessions guided by a research coordinator. A subset of Dataset I participants were assessed remotely via Microsoft Teams due to COVID-19 restrictions, completing the same tasks with the same stimuli. For Dataset II, participants completed tasks independently on their personal smartphones via the Winterlight iOS application.

#### Feature Extraction

For both datasets, audio recordings were transcribed using automated speech-to-text processing supplemented by human annotation, with examiner speech excluded. Speech and language features were extracted using the Winterlight platform (winterlightlabs.com), an automated pipeline described in detail in Tang et al. (2025). The platform uses Python-based acoustic and natural language processing libraries to extract features spanning acoustic/voice quality (via Praat/Parselmouth; Jadoul et al., 2018), morphosyntactic and syntactic complexity (via SpaCy, Honnibal et al., 2020; Stanford NLP parser, Chen & Manning, 2014), semantic coherence (via GloVe and FastText embeddings; Pennington et al., 2014; Bojanowski et al., 2017), lexical and psycholinguistic characteristics (based on published norms; Brysbaert et al., 2018; Kuperman et al., 2012; Warriner et al., 2013), discourse graph structure (Mota et al., 2012), and adjacent-word cosine similarity measures (Zhou et al., 2022). Task-specific features quantifying correctly identified picture description elements were also computed.

### Analytic Approach By Objective

**Objective A: Cross-Sectional Group Differences.**

#### Sample

**Dataset I (“ACES”)** was a cross-sectional sample used for Objective A. Recruitment was open to individuals with schizophrenia spectrum disorders, bipolar disorder, depressive disorders, and healthy volunteers, with a minimum age of 14 years. Only adults with depressive disorders (N=82) and healthy controls (N=80) were included in the present analysis (total N=162). Previous publications describe this dataset but do not focus on vocal biomarkers of depression (Tang et al., 2026).

#### Feature Set

We extracted 758 features per task from Dataset I. Mel-frequency cepstral coefficient features (168) and zero-crossing rate features (4) were excluded a priori owing to high dimensionality, limited interpretability, and sensitivity to recording conditions, leaving 586 candidate features for the analysis (Darch, 2008; Simmatis, 2023). For all tasks, features with >20% missing data or zero variance were removed. For the phonemic fluency task, morphosyntactic and semantic coherence features were additionally excluded. After filtering, 396 features remained for journaling, 428 for picture description, and 295 for phonemic fluency.

#### Statistical analysis

For each task, we first used two-sample Welch’s t-tests (two-tailed) to compare each feature between groups, with Cohen’s d (pooled SD) as the effect size. Then, to adjust for confounds, separate logistic regression models predicted diagnostic group from each z-scored feature (with features z-scored across the full combined sample), controlling for sex, assessment modality (virtual vs. in-person), and years of education (z-scored). Models were fitted via maximum likelihood with a logit link. Observations with missing values were excluded on a per-model basis and sample sizes ranged from 142 to 162 depending on feature-level missingness.

#### Multiple comparisons

FDR correction was applied within each task to the logistic regression p-values. Features surviving FDR correction at q <.05 were identified as candidate features for Objectives B and C.

### Objective B: Trajectories

#### Sample

**Dataset II (“ACES-MDD”)** was a longitudinal sample of adults with mood disorders used for Objectives B and C. Participants with bipolar disorder were excluded from analysis. Participants were followed for up to four timepoints, each approximately one month apart. The Patient Health Questionnaire-9 (PHQ-9) was administered at all timepoints (Kroenke, Spitzer, & Williams, 2001). For Objective B participants were required to have speech and PHQ-9 data at ≥2 timepoints (N=85). Participants completed a mean of 3.2 timepoints over a mean follow-up of 10±3.5 weeks. Mean baseline PHQ-9 was 13.7±7.0, indicating moderate depression severity which decreased by 3.7 points (SD = 5.5) on average from first to last assessment.

#### Feature Set

Significant FDR-corrected (q<0.05) speech features from Objective A were carried forward: 63 features for journaling, 16 for picture description, and 11 for phonemic fluency. For journaling and picture description, features were averaged across stimuli within each session. Features were z-scored across all observations.

#### Statistical Analysis

Following the analytic approach of Tang et al. (2025), random-intercept linear mixed models were fitted using the nlme package in R, estimated via maximum likelihood. Weeks from baseline were computed using session dates extracted from the speech data. Where session dates were unavailable, median study intervals were assigned based on timepoint (0, 3.6, 7.7, and 12.1 weeks for timepoints 1–4, respectively). Time was centered at baseline (week 0). The time structure was determined by comparing intercept-only, linear, and quadratic unconditional growth models; quadratic time was selected for all three tasks based on BIC. For each feature within each task, the following model was fitted: PHQ-9 ∼ speech feature + weeks + weeks² + speech feature × weeks + speech feature × weeks² + (1 | participant). P-values for the main effects, linear interactions, and quadratic interactions were FDR-corrected in separate families within each task. Model improvement was assessed by comparing BIC and AIC between each speech model and a time-only model (PHQ-9 ∼ weeks + weeks²) fitted to the same complete-case observations. Sex and age moderation were tested in separate models that added a speech feature × sex or speech feature × age interaction term. One participant who reported intersex status was included in all primary models but excluded from sex-stratified analyses.

#### Multiple comparisons

FDR correction was applied separately to main effects, linear interactions, quadratic interactions, sex interactions, and age interactions, each within task. We report FDR < .05 as the primary threshold and FDR < .10 as a secondary, exploratory threshold.

### Objective C. Treatment Prediction

#### Sample

From Dataset II, 64 participants had baseline PHQ-9 ≥ 10, indicating at least moderate severity, with at least one subsequent assessment. One was excluded due to intersex status, which was insufficient for stable estimation with sex as a predictor, and one was missing the phonemic fluency task. This yielded analytic samples of 63 for journaling, picture description, and the combined analysis, and 62 for phonemic fluency.

#### Feature Set

Only the features showing an FDR-corrected significant cross-sectional relationship with depression severity were included in this analysis (i.e., the 63 features from the Objective A results). Features were examined in combination and separately for each of the three tasks, resulting in four feature sets.

#### Statistical Analysis

The main predicted outcome was treatment response defined as ≥50% reduction in PHQ-9 total score from baseline to the last available follow-up assessment (Coley et al., 2020). For each of the four feature sets, three models were evaluated with different predictors: (a) clinical only (baseline PHQ-9, age, sex) (Coley 2021; Simon et al, 2024), (b) speech only, and (c) combined (clinical + speech). The prediction of treatment response was performed using elastic net logistic regression with 5-fold cross-validation, performed separately for each model. Elastic net logistic regression was used given the small sample size and high predictor-to-observation ratio. Penalty (λ) and mixing (α; 0 = ridge, 1 = lasso) parameters were tuned via grid search within the cross-validation procedure. Preprocessing included removal of zero– and near-zero-variance predictors, median imputation of missing feature values, and standardization of all numeric predictors, each applied within cross-validation folds to prevent data leakage. ROC-AUC was used as the primary metric for model evaluation, with incremental predictive value of each model (clinical, speech, combined) assessed by comparing fold-level AUC values using paired t-tests

#### Multiple comparisons

Model comparison paired t-test p-values were adjusted for multiple comparisons using the FDR correction, applied across comparisons within each feature set.

## Results

### Objective A

After FDR correction, 63 features survived at q < .05 for journaling, 16 for picture description, and 11 for phonemic fluency. In every task, the ten strongest effects, ranked by combining t-test effect sizes and covariate-adjusted regression coefficients, were exclusively voice quality measures: shimmer (cycle-to-cycle variability in vocal amplitude) and harmonics-to-noise ratio (HNR), the ratio of periodic to aperiodic energy in the voice signal, where lower values indicate a noisier, breathier voice. The depressed group showed higher shimmer and lower HNR across all three tasks. The three largest adjusted effects were |β| = 1.79 to 1.89 for journaling and 1.79 to 1.82 for picture description (all q < .001), and |β| = 1.49 to 1.66 for phonemic fluency (q = .004 to .007).

Eleven features, six shimmer variants and five HNR variants, survived correction in all three tasks, with consistent effect directions throughout (Figure 1). These variants are alternative algorithms for computing the same two underlying constructs and are highly intercorrelated. They are best interpreted as convergent estimates of shimmer and HNR rather than as eleven independent findings.

**Figure 1.**
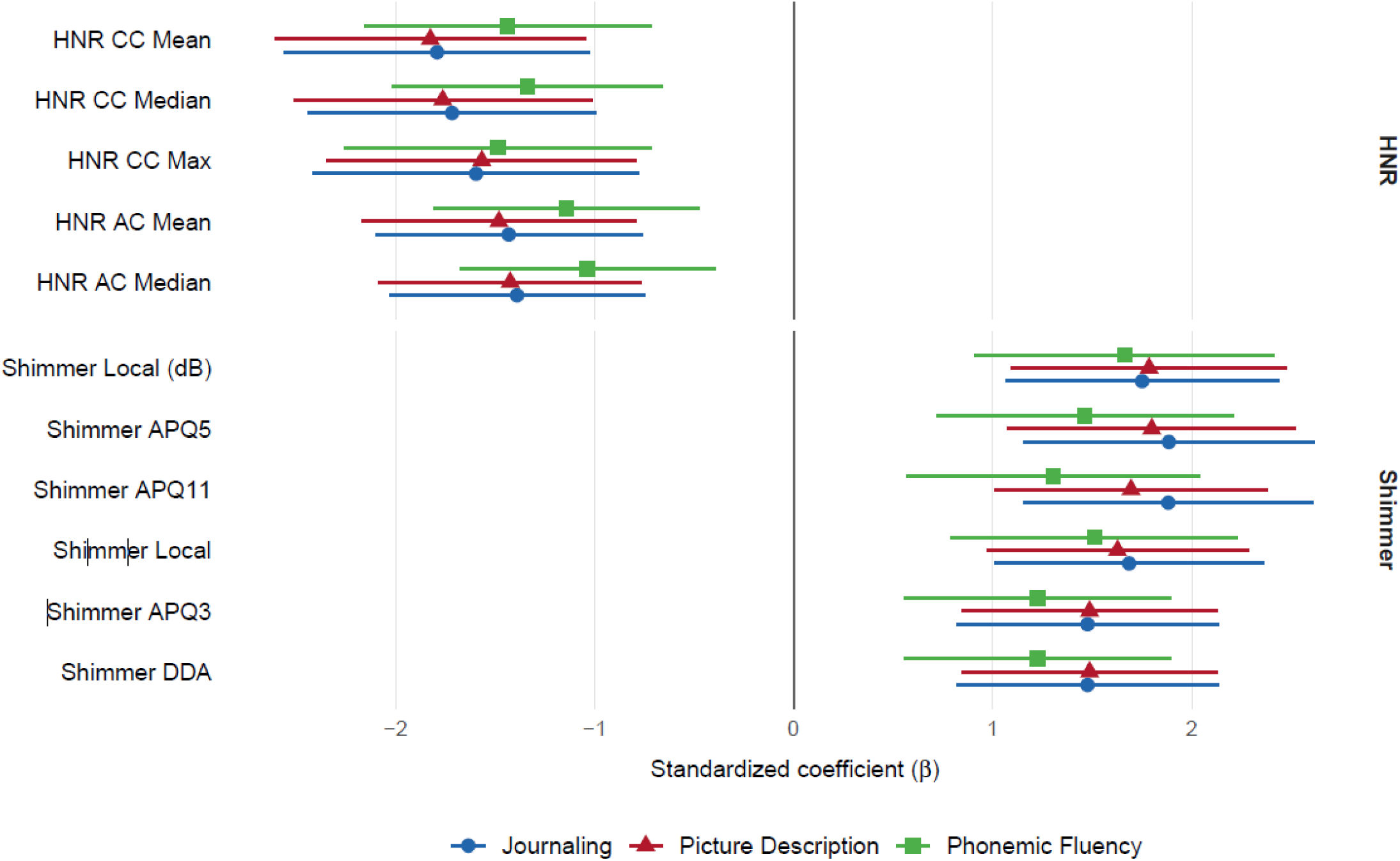
Covariate-adjusted associations between voice quality features and depression status across three speech tasks in the analysis of cross-sectional group differences. Standardized logistic regression coefficients and 95% confidence intervals for the 11 voice quality features surviving FDR correction (q < .05) in all three tasks. Positive values indicate higher scores in the depressed group; negative values indicate lower scores. All shimmer features were elevated, and all HNR features were reduced in the depressed group. Models controlled for sex, assessment modality, and education. N = 160 (journaling), N = 160 (picture description), N = 159 (phonemic fluency).

Journaling was the only task in which features beyond voice quality survived correction. Relative to controls, the depressed group produced longer utterances with greater syntactic complexity (more clauses and verb phrases per utterance, deeper parse trees), a higher proportion of subordinating conjunctions and auxiliary verbs, and longer total speaking duration. Lexical diversity was reduced on both indices examined (type-token ratio and Brunet’s Index, which is inversely scaled such that higher values indicate lower richness; both q < .02). Local semantic coherence was greater in the depressed group, reflected in smaller cosine distances between adjacent words on both coherence indices (q = .013 and .035).

### Objective B

PHQ-9 scores declined across the study period, with mean scores decreasing from approximately 13.7 at baseline to 9.1 at timepoint 4. Unconditional growth models indicated that a quadratic time structure best characterized PHQ-9 change in all three task samples (all quadratic terms(p < .001, ΔBIC = −12.5 to −13.0 relative to linear models), reflecting an initial steep decline in symptoms that decelerated over time. The intraclass correlation coefficient (ICC) was approximately .63 across tasks, indicating that 63% of PHQ-9 variance was between-person and 37% was within-person.

In phonemic fluency, one voice quality feature met the primary threshold (hnr_cc_max, FDR = .006). The remaining ten features fell at FDR .07 to .10 and were consistent in direction, with HNR measures negatively and shimmer measures positively associated with PHQ-9 (Figure 2). HNR features were negatively associated with PHQ-9: the strongest was hnr_cc_max (β = −1.43, SE = 0.41, t = −3.51, p = .0006, FDR = .006). indicating that a one standard deviation increase in maximum HNR was associated with a 1.43-point decrease in PHQ-9. Additional HNR features (hnr_cc_mean, hnr_ac_mean, hnr_cc_median, hnr_ac_median) showed consistent negative associations (β range: −0.79 to −0.87, FDR range: .073–.080). Shimmer features were positively associated with PHQ-9, indicating that greater amplitude perturbation (reflecting poorer voice quality) co-occurred with higher depression severity. These associations were stable across the treatment period: no speech × time interactions were significant (all FDR > .82), indicating that the voice quality–depression relationship did not change as symptoms improved.

**Figure 2.**
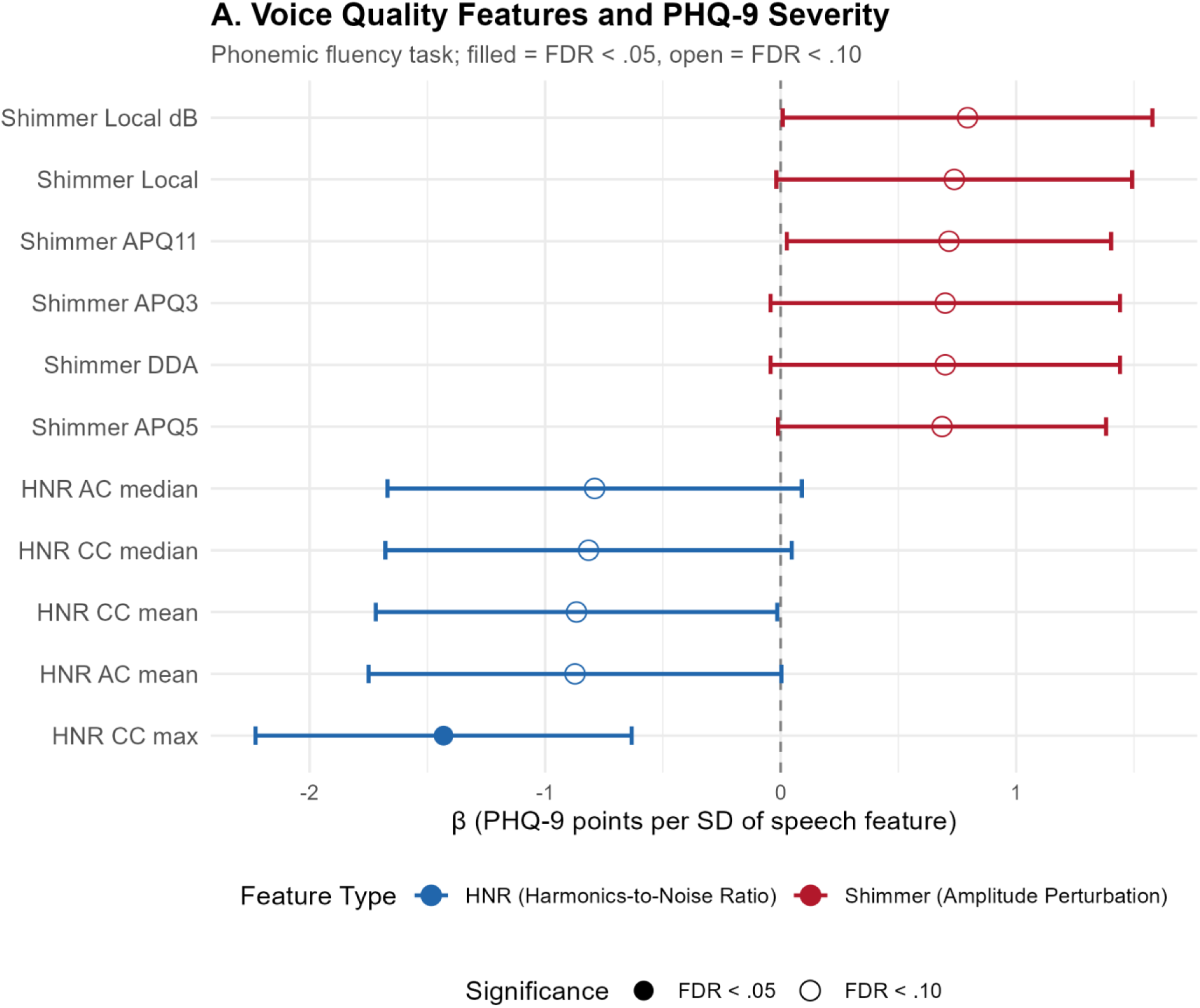
Forest plot of standardized regression coefficients (β) from random-intercept linear mixed models predicting PHQ-9 total score from time-varying voice quality features during the phonemic fluency task. Coefficients represent the expected change in PHQ-9 per standard deviation increase in each speech feature, adjusting for time and individual intercepts. HNR features were negatively associated with depression severity (blue), while shimmer features were positively associated (red). Filled circles indicate FDR-corrected p < .05; open circles indicate FDR-corrected p < .10. Error bars represent 95% confidence intervals.

Neither journaling nor picture description yielded FDR-significant associations between speech features and PHQ-9 (journaling: 0 of 63 features at FDR < .05; picture description: 0 of 16). In both tasks, the strongest uncorrected associations involved the same HNR variance features that were significant in phonemic fluency (journaling: hnr_cc_variance β = −1.02, p = .044; picture description: hnr_ac_variance β = −0.86, p = .059).

Sex moderated voice quality associations with PHQ-9 in picture description and phonemic fluency, but not journaling (Figure 3). Seven of 16 picture description features showed sex × speech interactions at FDR < .05, with a further eight at FDR < .10; 6 of 11 phonemic fluency features met the primary threshold (all FDR < .007), all of them shimmer variants. Simple slopes at baseline indicated that in females, higher shimmer and lower HNR were associated with higher PHQ-9, consistent with the cross-sectional direction of effect. In males, point estimates were reversed in all 13 features (|β| = 1.06 to 1.70), though no male slope was significant after correction and male subgroups comprised 20 participants per task. Sex did not moderate PHQ-9 trajectories in either task (all p > .20), indicating these interactions were not attributable to differential recovery. In journaling, 8 features showed sex interactions at uncorrected p < .05, none surviving correction. Age did not moderate speech associations in any task.

**Figure 3.**
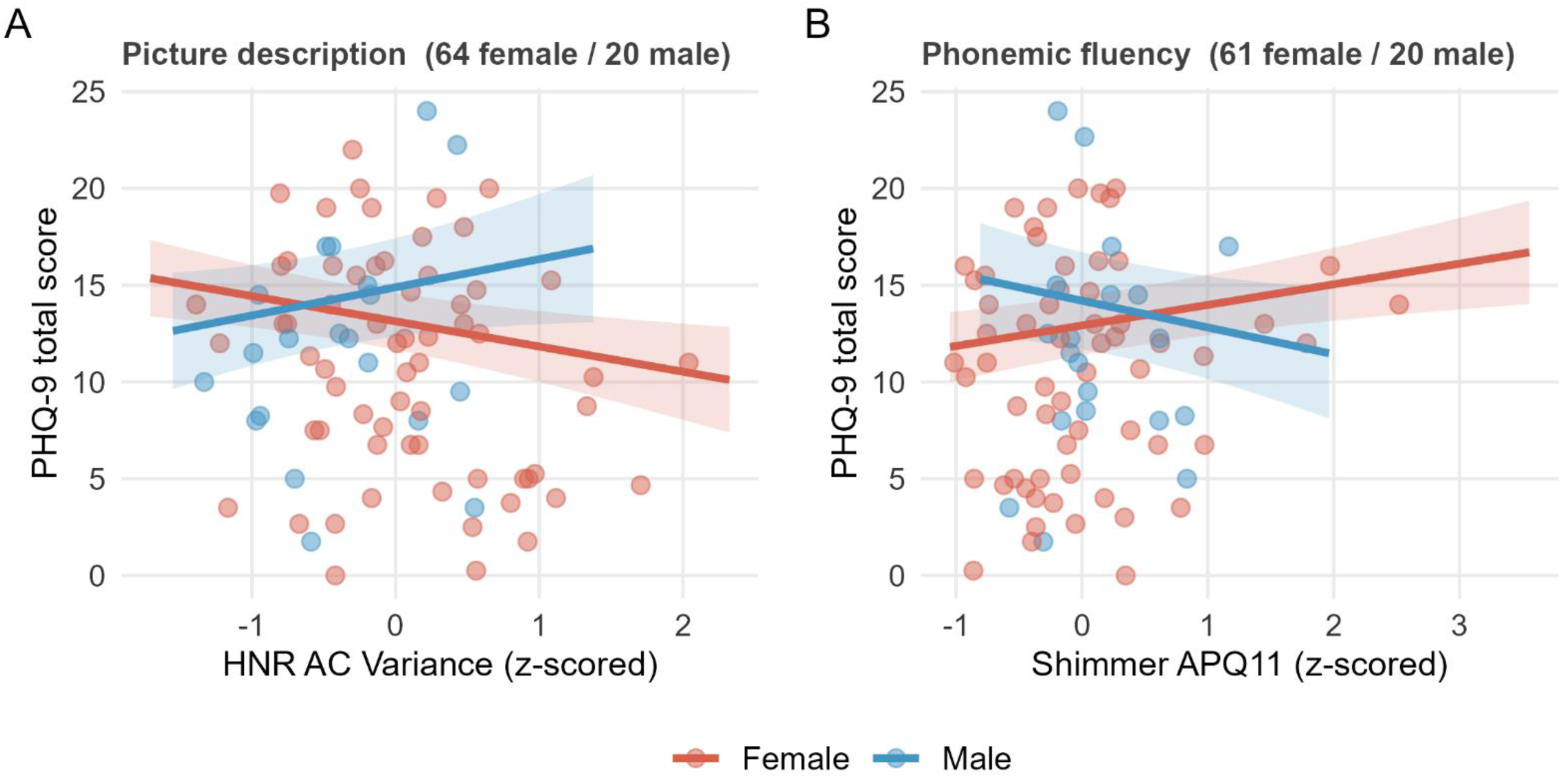
Sex moderation of voice quality associations with depression severity. Model-implied associations between PHQ-9 and (A) HNR AC variance during picture description and (B) shimmer APQ11 during phonemic fluency, estimated separately by sex from random-intercept linear mixed models adjusting for time. Slopes are evaluated at baseline. Shaded bands are 95% confidence intervals for the fixed effects; points are participant means, which scatter beyond these bands. Sex × speech interactions: (A) β = −2.76, SE = 0.82, FDR = .006; (B) β = 2.45, SE = 0.72, FDR = .007. Male estimates derive from a smaller subgroup (n = 20 in both tasks; female n = 64 and 61, respectively) and individual male slopes were not significant after correction. Shimmer and HNR variants are computed by closely related algorithms and are highly intercorrelated; the features shown are representative of two constructs rather than independent findings. HNR, harmonics-to-noise ratio; APQ, amplitude perturbation quotient.

### Objective C

Cross-validated AUCs ranged from 0.653 to 0.730 across the twelve models (Table 2). The highest discrimination came from combining speech features across all tasks with clinical features (AUC = 0.730 [0.639, 0.820]; sensitivity 0.681, specificity 0.742). Among single-task models, phonemic fluency speech features performed best (AUC = 0.727 [0.662, 0.792]), paralleling the Objective B finding that phonemic fluency was the only task yielding longitudinal associations with symptom severity. Journaling and picture description speech-only models performed less well (AUC = 0.653 and 0.657). Clinical-only models achieved AUC = 0.700 to 0.715.

**Table 2:** Cross-validated prediction of treatment response using clinical, speech, and combined predictor sets. Treatment response was defined as ≥50% reduction in PHQ-9 from baseline to last available follow-up. AUC, sensitivity, and specificity are from 5-fold cross-validation repeated 3 times, stratified by treatment response. Sensitivity and specificity are reported at the Youden-optimized threshold. Clinical features were baseline PHQ-9, age, and sex. Speech predictors were FDR-surviving features from Objective A. Elastic net logistic regression was used for all models. N = 63 for all analyses except phonemic fluency (N = 62).

| Task | Predictor Set | AUC | 95% CI | p value | Sensitivity | Specificity |
| --- | --- | --- | --- | --- | --- | --- |
| All Tasks Combined | Clinical Only | 0.715 | [0.612, 0.818] | <.001 | 0.58 | 0.758 |
| All Tasks Combined | Speech Only | 0.704 | [0.626, 0.781] | <.001 | 0.594 | 0.708 |
| All Tasks Combined | Clinical + Speech | 0.730 | [0.639, 0.82] | <.001 | 0.681 | 0.742 |
| Journaling | Clinical Only | 0.715 | [0.612, 0.818] | <.001 | 0.58 | 0.758 |
| Journaling | Speech Only | 0.653 | [0.588, 0.719] | <.001 | 0.652 | 0.625 |
| Journaling | Clinical + Speech | 0.690 | [0.604, 0.776] | <.001 | 0.725 | 0.708 |
| Phonemic Fluency | Clinical Only | 0.700 | [0.596, 0.804] | <.001 | 0.565 | 0.778 |
| Phonemic Fluency | Speech Only | 0.727 | [0.662, 0.792] | <.001 | 0.594 | 0.726 |
| Phonemic Fluency | Clinical + Speech | 0.691 | [0.624, 0.759] | <.001 | 0.638 | 0.65 |
| Picture Description | Clinical Only | 0.715 | [0.612, 0.818] | <.001 | 0.58 | 0.758 |
| Picture Description | Speech Only | 0.657 | [0.584, 0.729] | <.001 | 0.928 | 0.292 |
| Picture Description | Clinical + Speech | 0.726 | [0.652, 0.801] | <.001 | 0.855 | 0.517 |

No paired comparison of fold-level AUCs between predictor configurations reached significance after correction (all FDR-corrected p > .54). Speech features therefore did not demonstrate incremental predictive value over clinical variables alone, nor the reverse.

Voice quality features were the most consistently retained predictors, appearing in every task-level model with selection in 100% of cross-validation folds (Supplemental Table 1). Coefficient signs for individual shimmer and HNR variants should not be interpreted directionally. Because these variants are computed by closely related algorithms and are highly intercorrelated, penalized regression distributes weight across them in ways that can produce opposing signs within a correlated set, as observed here for shimmer in the phonemic fluency model. The stable finding is that voice quality features as a family were retained across tasks and folds, not the direction of any individual variant.

## Discussion

This study examined whether speech features distinguish adults with depression from healthy controls, track with symptom severity during naturalistic treatment, and predict treatment response. Features were identified in a cross-sectional sample and then carried forward to a separate, non-overlapping longitudinal sample, providing a test of whether cross-sectional group differences generalize to within-cohort clinical change. Across objectives, voice quality features, specifically shimmer and HNR, emerged as the most consistent speech correlates of depression. Eleven voice quality features survived FDR correction in all three tasks cross-sectionally. In the longitudinal sample, phonemic fluency voice quality features were consistent in direction across all 11 features, though only maximum HNR met the primary significance threshold. In the prediction sample, phonemic fluency features achieved the highest single-task discrimination for treatment response (AUC = 0.727), comparable to clinical features alone.Voice quality features distinguished depressed individuals from healthy controls. Eleven voice quality features survived FDR correction across all three tasks, with consistent effect directions: higher shimmer (greater amplitude perturbation) and lower HNR (noisier vocal signal) in the depressed group. These associations held after adjustment for sex, assessment modality, and education. The consistency of voice quality findings across tasks with distinct cognitive and linguistic demands suggests that voice quality differences in depression are not an artifact of task-specific demands but may reflect a more general property of vocal production. While prior studies have emphasized timing and prosodic features as the primary speech correlates of depression (Cantor-Cutiva et al., 2026, Yamamoto 2020), voice quality features have received comparatively little attention. The present findings suggest that voice quality may warrant greater emphasis in future research.

Beyond voice quality, journaling was the only task in which linguistic features survived FDR correction. Relative to controls, the depressed group produced longer, more syntactically complex utterances with lower lexical diversity and greater local semantic coherence, reflected in smaller semantic distances between adjacent words. The verbosity and reduced lexical richness are consistent with prior reports (Menne et al., 2024), and together with greater local coherence suggest repetitive, semantically constrained speech drawn from a narrower conceptual range. The syntactic complexity finding is less expected, running counter to the classical characterization of depressed speech as sparse and impoverished (Kendler, 2017). These features did not carry forward to the longitudinal sample, though the journaling composite differed between datasets. In the longitudinal sample it averaged responses to an open-ended prompt with a prompt eliciting positive expectations for the coming week, a more constrained format likely to narrow the range of linguistic output. This blending may have diluted signal present in the open-ended responses alone. Whether these linguistic features are genuinely state-insensitive or were obscured by task composition cannot be resolved with the data presented here.

The emergence of phonemic fluency as the task yielding longitudinal effects parallels the prediction results and may reflect the task’s unique properties. One minute of constrained single-word production minimizes linguistic variability and isolates acoustic-vocal signals, providing a cleaner window into voice quality than tasks requiring complex language production, such as journaling tasks. This has practical implications: if voice quality is the primary signal of interest, brief constrained tasks may be more efficient assessment vehicles than longer open-ended speech samples. In the longitudinal sample, phonemic fluency was the only task to show associations with PHQ-9 severity after FDR correction. The strongest association was for hnr_cc_max. One standard deviation increase in maximum HNR was associated with a 1.43-point decrease in PHQ-9. On a scale where a 5-point change is considered the minimal clinically important difference (Kroenke, 2001; Löwe 2004), this represents a modest but meaningful shift. These associations were driven substantially by between-person differences. Approximately 63% of PHQ-9 variance was between-person, meaning participants with chronically higher symptom levels also tended to have chronically poorer voice quality. This does not establish that voice quality worsens within an individual as symptoms worsen. Models of the present form estimate a mixture of between– and within-person association and cannot separate them. Partitioning these sources, for instance by modeling within-subject deviation scores or correlating change scores, would provide a more direct test of whether voice quality functions as a state marker suitable for treatment monitoring.

Several design features of the study may have limited our ability to detect within-person effects. The sample was modest in size, participants contributed 2-4 timepoints each. Importantly, participants were recruited at varying points in their treatment rather than at treatment initiation, with heterogenous pharmacological and therapeutic exposures, Some participants may have already experienced substantial improvement before their first recorded timepoint, while others may be early in their treatment. This heterogeneity compresses the observable range of within-person symptom change, reducing power to detect features that covary with clinical improvement. A study recruiting at treatment initiation, with more frequent assessments during the period of most rapid symptom change, would provide a stronger test of whether voice quality features are sensitive to within-person clinical trajectories.

An unanticipated finding was that sex significantly moderated the relationship between voice quality features and depression severity in both picture description and phonemic fluency. One interpretation is that voice quality is a more sensitive marker of depression in females. Male and female voices differ in vocal fold size and fundamental frequency, which affects how perturbation measures such as shimmer and HNR behave (Amir, 2004; Kaczmarek-Majer, 2024; Titze, 1989) . In the present sample, male slopes ran opposite to the expected direction, but they were not significantly different from zero and came from only 20 men per task. A possible explanation is that the male estimates are imprecise rather than genuinely reversed. Determining whether voice quality is simply less sensitive in males, or behaves differently altogether, will require a sample with more male participants..

Speech-only models achieved cross-validated AUCs ranging from 0.653 to 0.727, with phonemic fluency voice quality features yielding the highest discrimination (AUC = 0.727). These estimates were comparable to clinical-only models based on baseline PHQ-9, age, and sex (AUC = 0.700–0.715). However, no pairwise comparison between predictor configurations reached statistical significance after correction for multiple comparisons, precluding conclusions about incremental predictive value. The wide confidence intervals and high variance inherent to repeated cross-validation in samples of this size mean that the present analyses are best interpreted as establishing feasibility and preliminary effect sizes rather than definitive evidence of predictive utility. Still, the pattern of results was nonetheless informative. Voice quality features were consistently retained by regularized models across tasks and cross-validation folds with stable directional effects. The finding that a one-minute phonemic fluency recording achieves discrimination comparable to clinical variables has practical implications: such recordings could be collected passively or semi-passively in clinical settings at minimal cost, potentially supplementing clinician judgment in treatment planning.

### Strengths

This study has several methodological strengths that distinguish it from much of the existing literature. First, the use of two separate samples: one which establishes cross-sectional group differences, and another that tests longitudinal prediction, provides a more rigorous test of generalizability than single-sample analyses. Second, the systematic comparison of features across three speech tasks within the same participants allows evaluation of task-specificity, an underexplored dimension with direct implications for clinical deployment. Third, the analytic approach prioritized interpretability: individual feature-level tests with FDR correction, rather than black-box classifiers, enable direct identification of which speech properties are most informative and facilitate replication.

### Limitations

Both datasets were drawn from a single community mental health center using convenience sampling. While the use of nonoverlapping datasets for cross-sectional and longitudinal evaluation is a strength, samples were modest in size, diagnoses were chart-derived rather than based on structured interviews, and participants were recruited at varying points in their treatment trajectories with heterogeneous pharmacological and therapeutic exposures. While this design may enhance ecologic validity, it may also introduce noise. Recording conditions varied within and across datasets; although assessment modality was covaried in cross-sectional analyses, residual confounding from microphone hardware, environment, and audio compression cannot be excluded, particularly for acoustic features known to be sensitive to recording conditions.

### Conclusions

Voice quality features, particularly shimmer and HNR, emerged as the most robust and consistent speech correlates of depression across cross-sectional, longitudinal, and predictive analyses and across multiple elicitation tasks. These features are computationally simple to extract, require only brief speech samples, and showed preliminary evidence of clinical utility in predicting treatment response during naturalistic care. The finding that these associations are stronger in females generates a testable hypothesis for future research in sex-specific contexts such as perinatal depression. Larger, prospectively designed studies with standardized recording conditions, structured diagnostic assessments, and recruitment at treatment initiation are needed to establish whether voice quality features can serve as scalable, objective biomarkers for depression monitoring and treatment planning.

## Data Availability

All data produced in the present study are available upon reasonable request to the authors.

## Acknowledgements

Sarah Berretta, Sophia Deligiannidis, Hannah Contreras

---

**Supplemental Table 1:**
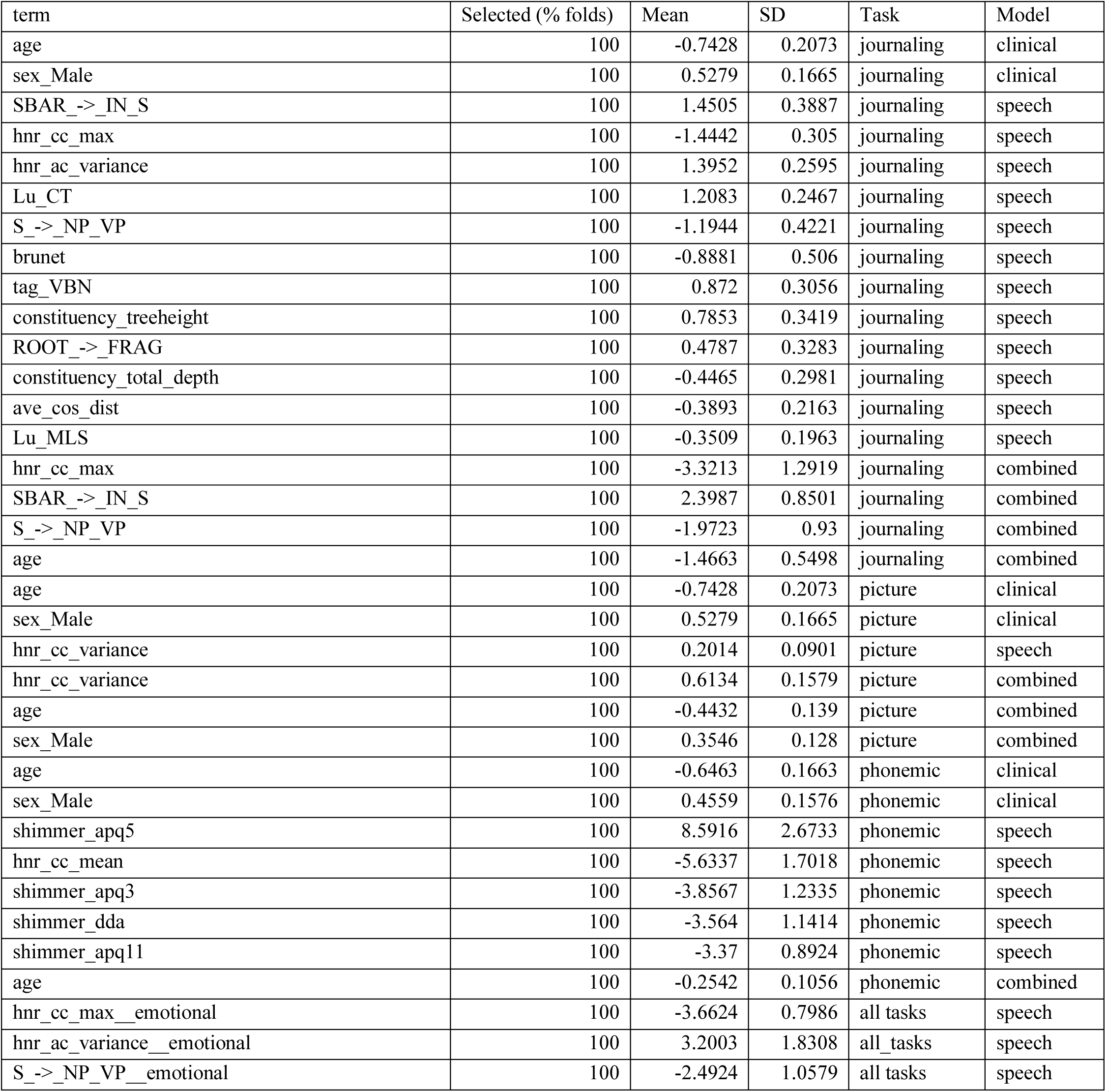
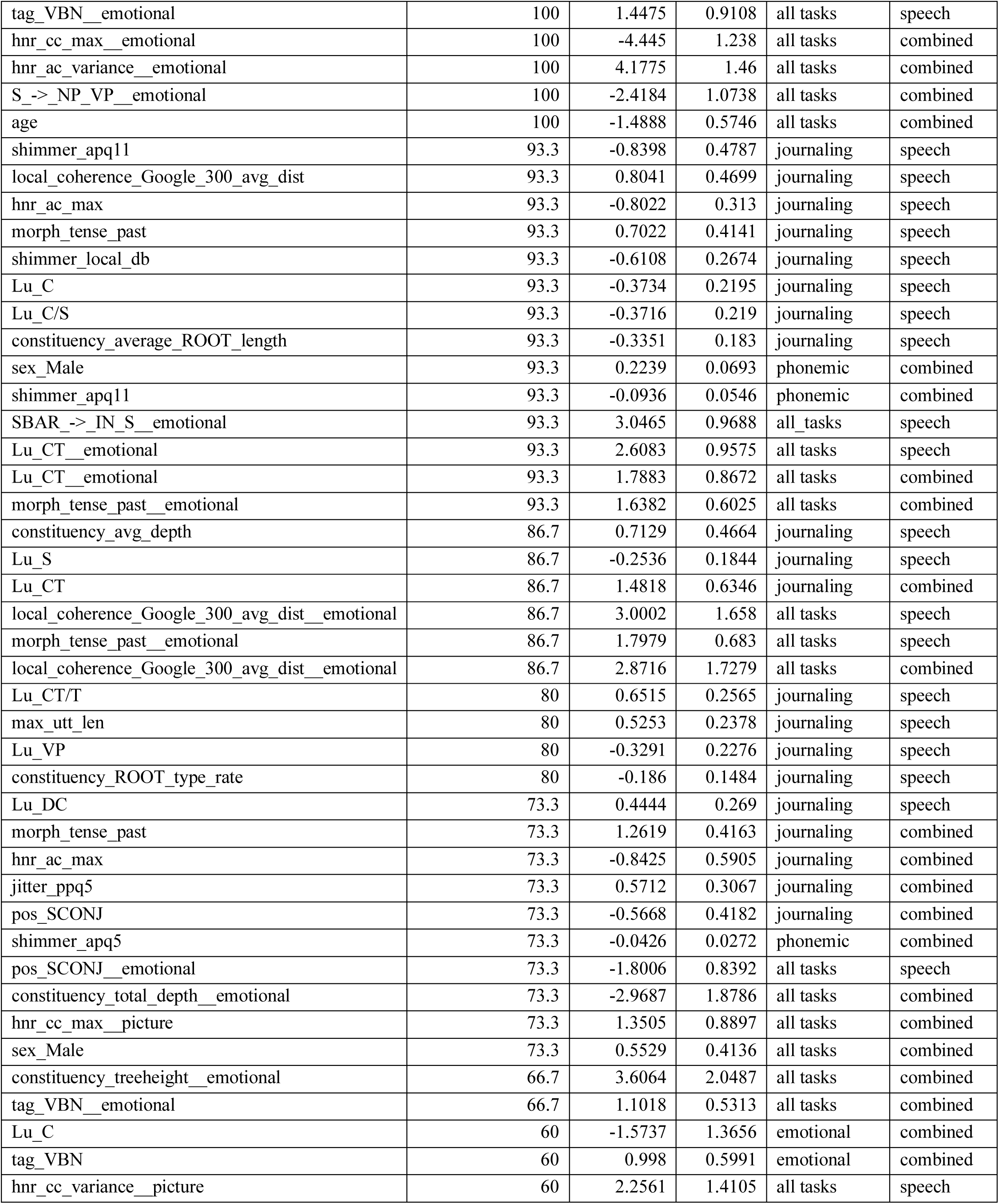
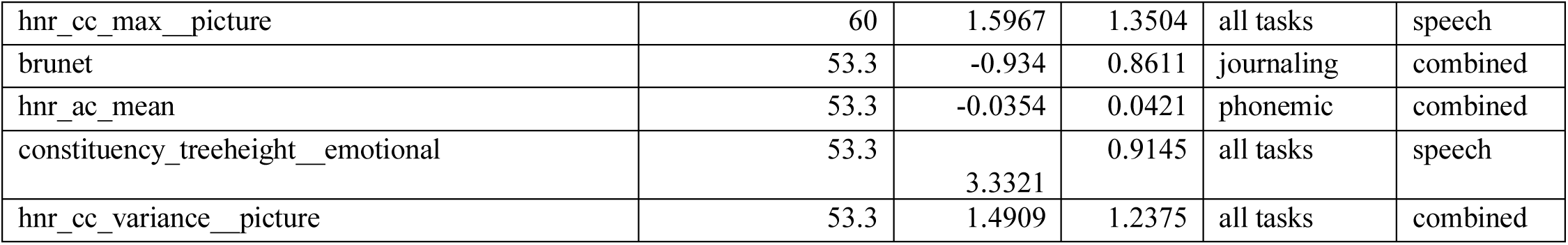
Elastic net coefficient stability across cross-validation folds for treatment response prediction models. Features selected in >50% of cross-validation folds are shown. “Selected (% folds)” indicates the percentage of folds in which the feature received a nonzero coefficient. Mean β and SD are computed across folds in which the feature was selected. Features are grouped by task and model (clinical only, speech only, combined) and sorted by selection frequency within each group. For the all-tasks-combined models, feature names are suffixed with the originating task.

## References

1. Amir O, Biron-Shental T. The impact of hormonal fluctuations on female vocal folds. Curr Opin Otolaryngol Head Neck Surg. 2004;12(3):180–4.

2. Bojanowski P, Grave E, Joulin A, Mikolov T. Enriching word vectors with subword information. Trans Assoc Comput Linguist. 2017;5:135–46.

3. Briganti G, Lechien JR. Speech and voice quality as digital biomarkers in depression: a systematic review. J Voice. 2025. doi:10.1016/j.jvoice.2025.05.002.

4. Brysbaert M, Mandera P, McCormick SF, Keuleers E. Word prevalence norms for 62,000 English lemmas. Behav Res Methods. 2019;51(2):467–79.

5. Cai H, Jin Y, Liu S, Zhang Q, Zhang L, Cheung T, et al. Prevalence of suicidal ideation and planning in patients with major depressive disorder: a meta-analysis of observation studies. J Affect Disord. 2021;293:148–58.

6. Cai H, Xie XM, Zhang Q, Cui X, Lin JX, Sim K, et al. Prevalence of suicidality in major depressive disorder: a systematic review and meta-analysis of comparative studies. Front Psychiatry. 2021;12:690130.

7. Cantor-Cutiva LC, Carrillo-González A, Yousef AM, Berardi M, Hunter EJ. The role of voice acoustics in depression assessment: findings from bibliometric analysis, literature review, and meta-analysis. Depress Anxiety. 2026;2026:5592230.

8. Chen D, Manning CD. A fast and accurate dependency parser using neural networks. In: Proceedings of the 2014 Conference on Empirical Methods in Natural Language Processing. Doha, Qatar: Association for Computational Linguistics; 2014. p.740–50.

9. Coley RY, Boggs JM, Beck A, Hartzler AL, Simon GE. Defining success in measurement-based care for depression: a comparison of common metrics. Psychiatr Serv. 2020;71(4):312–8.

10. Coley RY, Boggs JM, Beck A, Simon GE. Predicting outcomes of psychotherapy for depression with electronic health record data. J Affect Disord Rep. 2021;6:100198.

11. Darch J, Milner B, Vaseghi S. Analysis and prediction of acoustic speech features from mel-frequency cepstral coefficients in distributed speech recognition architectures. J Acoust Soc Am. 2008;124(6):3989–4000.

12. Dong M, Zeng LN, Lu L, Li XH, Ungvari GS, Ng CH, et al. Prevalence of suicide attempt in individuals with major depressive disorder: a meta-analysis of observational surveys. Psychol Med. 2019;49(10):1691–704.

13. Ferreira F, Barker M. Perceptual clauses as units of production in visual descriptions. Top Cogn Sci. 2025;17(4):868–76.

14. Garcia ME, Hinton L, Neuhaus J, Feldman M, Livaudais-Toman J, Karliner LS. Equitability of depression screening after implementation of general adult screening in primary care. JAMA Netw Open. 2022;5(8):e2227658.

15. Honnibal M, Montani I, Van Landeghem S, Boyd A. spaCy: industrial-strength natural language processing in Python. 2020.

16. Jadoul Y, Thompson B, de Boer B. Introducing Parselmouth: a Python interface to Praat. J Phonetics. 2018;71:1–15.

17. Kaczmarek-Majer K, Dominiak M, Antosik AZ, Hryniewicz O, Kamińska O, Opara K, et al. Acoustic features from speech as markers of depressive and manic symptoms in bipolar disorder: a prospective study. Acta Psychiatr Scand. 2025;151(3):358–74.

18. Kendler KS. The genealogy of major depression: symptoms and signs of melancholia from 1880 to 1900. Mol Psychiatry. 2017;22(11):1539–53.

19. Kroenke K, Spitzer RL, Williams JBW. The PHQ-9: validity of a brief depression severity measure. J Gen Intern Med. 2001;16(9):606–13.

20. Kuperman V, Stadthagen-Gonzalez H, Brysbaert M. Age-of-acquisition ratings for 30,000 English words. Behav Res Methods. 2012;44(4):978–90.

21. Löwe, B., Unützer, J., Callahan, C. M., Perkins, A. J., & Kroenke, K. (2004). Monitoring depression treatment outcomes with the patient health questionnaire-9. Medical care, 42(12), 1194–1201.

22. Mao K, Wu Y, Chen J. A systematic review on automated clinical depression diagnosis. npj Ment Health Res. 2023;2(1):20.

23. Maran PL, Braquehais MD, Vlaic A, Alonzo-Castillo MT, Vendrell-Serres J, Ramos-Quiroga JA, et al. Performance of automatic speech analysis in detecting depression: systematic review and meta-analysis. JMIR Ment Health. 2025;12:e67802.

24. Menne F, Dörr F, Schräder J, et al. The voice of depression: speech features as biomarkers for major depressive disorder. BMC Psychiatry. 2024;24(1):794. doi:10.1186/s12888-024-06253-6

25. Morgan CD, Murray HA. A method for investigating fantasies: the Thematic Apperception Test. Arch Neurol Psychiatry. 1935;34(2):289–306.

26. Mota NB, Vasconcelos NAP, Lemos N, Pieretti AC, Kinouchi O, Cecchi GA, et al. Speech graphs provide a quantitative measure of thought disorder in psychosis. PLoS One. 2012;7(4):e34928.

27. Mundt JC, Snyder PJ, Cannizzaro MS, Chappie K, Geralts DS. Voice acoustic measures of depression severity and treatment response collected via interactive voice response (IVR) technology. J Neurolinguistics. 2007;20(1):50–64.

28. Mundt JC, Vogel AP, Feltner DE, Lenderking WR. Vocal acoustic biomarkers of depression severity and treatment response. Biol Psychiatry. 2012;72(7):580–7.

29. Patterson J. Controlled oral word association test. In: Kreutzer JS, DeLuca J, Caplan B, editors. Encyclopedia of Clinical Neuropsychology. New York, NY: Springer; 2011.

30. Pennington J, Socher R, Manning CD. GloVe: global vectors for word representation. In: Proceedings of the 2014 Conference on Empirical Methods in Natural Language Processing. Doha, Qatar: Association for Computational Linguistics; 2014. p.1532–43.

31. Rorschach H. Psychodiagnostik: Methodik und Ergebnisse eines wahrnehmungsdiagnostischen Experiments. Bern: Bircher; 1921.

32. Roth C, Boston Diagnostic Aphasia Examination, Kreutzer/DeLuca/Caplan eds., Encyclopedia of Clinical Neuropsychology, Springer 2011.

33. Rush AJ, Trivedi MH, Wisniewski SR, Nierenberg AA, Stewart JW, Warden D, et al. Acute and longer-term outcomes in depressed outpatients requiring one or several treatment steps: a STAR*D report. Am J Psychiatry. 2006;163(11):1905–17.

34. Simmatis LER, Robin J, Pommée T, McKinlay S, Sran R, Taati N, et al. Validation of automated pipeline for the assessment of a motor speech disorder in amyotrophic lateral sclerosis (ALS). Digit Health. 2023;9:20552076231219102.

35. Simon GE, Cruz M, Boggs JM, Beck A, Shortreed SM, Coley RY. Predicting outcomes of antidepressant treatment in community practice settings. Psychiatr Serv. 2024;75(5):419–26.

36. Simon GE, Moise N, Mohr DC. Management of depression in adults: a review. JAMA. 2024;332(2):141–52.

37. Tang S, Partlan R, Nikzad A, Richardson A, Cho S, Cong Y, et al. An automated integrative pipeline for extracting quantitative, objective speech and language features relevant to mental health. ISCTM 2025 Autumn Conference. 2025.

38. Tang SX, Li J, Brosch K, et al. ACES: Ascertaining diagnosis Classification with Elicited Speech in Individuals with Heterogeneous, Comorbid Psychiatric Disorders. Psychiatry Research. 2026:117389. doi:10.1016/j.psychres.2026.117389

39. Titze IR. Physiologic and acoustic differences between male and female voices. J Acoust Soc Am. 1989;85(4):1699–707.

40. Warriner AB, Kuperman V, Brysbaert M. Norms of valence, arousal, and dominance for 13,915 English lemmas. Behav Res Methods. 2013;45(4):1191–207.

41. Yamamoto M, Takamiya A, Sawada K, Yoshimura M, Kitazawa M, Liang KC, et al. Using speech recognition technology to investigate the association between timing-related speech features and depression severity. PLoS One. 2020;15(9):e0238726.

42. Yawatkar V, Chow HM, Usler E. Automatic temporal analysis of speech: a quick and objective pipeline for the assessment of overt stuttering. Behav Res. 2025;57:228.

43. Zallar LJ, Dupont MB. Brain and blood biomarkers of major depressive disorder: a systematic review. JAMA Psychiatry. 2026;83(4):414–25.

44. Zhou J, Zhang Y, He S, Xu S, Sun Q, Zhao T, et al. Accelerated global burden of depressive disorders during the COVID-19 pandemic from 2019 to 2021. Sci Rep. 2025;15(1):9529.

45. Zhou K, Ethayarajh K, Card D, Jurafsky D. Problems with cosine as a measure of embedding similarity for high frequency words. In: Muresan S, Nakov P, Villavicencio A, editors. Proceedings of the 60th Annual Meeting of the Association for Computational Linguistics (Volume 2: Short Papers). Dublin, Ireland: Association for Computational Linguistics; 2022. p.401–23.

